# Do smoking and alcohol behaviours influence risk of type 2 diabetes? A Mendelian randomisation study

**DOI:** 10.1101/2024.07.26.24311054

**Authors:** Zoe E. Reed, Hannah M. Sallis, Rebecca C. Richmond, Angela S. Attwood, Deborah A. Lawlor, Marcus R. Munafò

## Abstract

**Background:** Previous studies suggest that smoking and higher alcohol consumption are both associated with greater risk of type 2 diabetes (T2D). However, studies examining whether these associations reflect causal relationships are limited and do not consider continuous glycaemic traits. The aim of the study was to determine whether there are causal effects of smoking and alcohol consumption on T2D risk and related glycaemic traits.

**Methods and Findings:** We conducted both two-sample and one-sample MR to examine the effects of lifetime smoking index (LSI) and alcoholic drinks per week on T2D and continuous traits (fasting glucose, fasting insulin and glycated haemoglobin, HbA1c). For two-sample MR we used results from genome-wide association studies (GWAS) of LSI (N=462,690), alcohol consumption (N=941,280), T2D (N= 148,726 cases and 965,732 controls) and continuous traits (N=149,289 to 209,605). We used inverse variance weighting (IVW) for our main analyses and conducted several sensitivity analyses to explore violation of MR assumptions. We compared two-sample MR to one-sample MR results for alcohol effects on T2D and HbA1c in UK Biobank (N=336,984). Only these analyses were conducted to avoid sample overlap and due to data availability. The main IVW two-sample MR results suggested possible causal effects of higher LSI on T2D risk (OR per 1SD higher LSI=1.42, 95% CI=1.22 to 1.64); however, sensitivity analyses did not consistently support this finding, and there was evidence of potential horizontal pleiotropy. There was no robust evidence that higher drinks per week influenced risk of T2D from our main IVW two-sample MR analyses (OR per 1 SD higher log-transformed drinks per week=1.04, 95% CI=0.40 to 2.65), despite evidence of causal effects on higher fasting glucose (difference in mean fasting glucose in mmol/l per 1SD higher log-transformed drinks per week=0.34, 95% CI=0.09 to 0.59). One-sample MR results suggested a possible causal effect of higher drinks per week on T2D risk (OR per 1 SD higher log-transformed drinks per week=1.71, 95% CI: 1.24 to 2.36), but in contrast, lower HbA1c levels (difference in mean SD of log transformed HbA1c (mol/mol) per 1 SD higher log-transformed drinks per week=-0.07, 95% CI: -0.11 to -0.02). Key limitations include limited generalisability of results due to analyses being conducted in European populations, and potential selection bias in UK Biobank influencing results.

**Conclusion:** Our results suggest effective public health interventions to prevent and/or reduce smoking and alcohol consumption are unlikely to reduce the prevalence of T2D.

## Introduction

Type 2 diabetes (T2D) is a common chronic condition that is known to increase risk of macro- and micro- vascular atherosclerotic diseases (1–4). Over the last 30 years, prevalence and incidence of T2D has increased markedly (5), and the age at which it is first diagnosed has decreased (6). These changes are largely thought to be driven by the global obesity epidemic (7), including increased testing for T2D in people who are obese. Whilst higher body mass index (BMI) is an established causal risk factor for T2D (8), other risk factors have been proposed.

Both smoking and alcohol have been suggested as potential risk factors that may causally affect T2D. Observationally, smoking is associated with higher risk, with heavier smokers having the greatest risk (9–12). Additionally, former smokers seem to have a higher risk for T2D compared to never smokers, with risk lowering over time since they quit (10,13). For alcohol consumption, there is a long history of observational studies suggesting a J-shaped association with cardiovascular diseases (14–16), with some studies finding evidence of a similar pattern with T2D (17,18). This slightly higher risk among those who report no alcohol consumption may be an artefact, for example due to misreporting, or because some people stop drinking (or never start) for health reasons (11,19). Despite this apparently higher risk at lower levels, across most of the distribution, higher alcohol consumption is associated with higher risk for T2D. The associations of smoking and alcohol with T2D might be causal or they might be influenced by confounding due to limited adjustment for socioeconomic position and related factors or only partially accounting for these in previous studies. In this study we aim to determine whether the relationships between smoking and alcohol and T2D are causal.

Mendelian randomisation (MR) is a causal inference method, which commonly uses genetic variants, typically single nucleotide polymorphisms (SNPs), as instrumental variables (IVs) for the exposure of interest. MR is less prone to confounding by socioeconomic, environmental and behavioural characteristics, or reverse causation than conventional observational studies (20). It can be biased by violation of the core assumptions underlying MR (Box 1).

#### Box 1. Core assumptions of Mendelian Randomisation

1) *The genetic IV is robustly associated with the exposure of interest in the relevant population (**relevance**)*.

May cause biased results if there are weak instruments (i.e., a statistically weak association of the genetic instrument with the exposure, which would bias results towards the null in two-sample MR and towards the confounded association in one-sample MR. In this study, comparing results from two- and one-sample MR for some effects is useful as, if both give consistent results this gives us greater confidence that neither have weak instrument bias. Population relevance is particularly important in two-sample MR, where it is important to ensure that the underlying population is the same in both samples and consistent with the population that we want to make inference to. Therefore, in this study we restricted analyses to include results from the genetic instrument-exposure and genetic instrument-outcome associations in white European adult populations only.

2) *There is no confounding between the genetic IV and the outcome (**independence**)*.

May be violated if there is population stratification, assortative mating, or confounding by dynastic factors. In this study we tried to minimise population stratification by only including results / data from participants of European ancestry and adjusting for principal components in our one-sample MR analyses.

3) *The genetic IV is only associated with the outcome via the exposure, and there are no direct effects of the genetic IV on the outcome (**exclusion restriction**)*.

May be violated if there is horizontal pleiotropy (i.e. the genetic variants influence risk factors for the outcome, independently of the exposure of interest. In this study we explored the likelihood of unbalanced horizontal pleiotropy influencing our main two-sample MR IVW results through comparing those results to results from several pleiotropy robust sensitivity analyses.

Two previous two-sample MR studies have found evidence for a potential causal effect of smoking initiation (21) and lifetime smoking (22) on risk of T2D. The former study used data from a genome wide association study (GWAS) of diabetes with 74,124 cases and 824,006 controls and focused on smoking initiation, which does not capture other important smoking related behaviours, like smoking duration and intensity. The latter study used data from the same diabetes GWAS and was an atlas study that examined many outcomes, and therefore did not focus on this relationship, nor did it consider continuous traits related to diabetes.

We identified two previous MR studies examining alcohol and T2D. The first reported no effect. It used one SNP from the alcohol dehydrogenase 1B gene which encodes an enzyme involved in alcohol metabolism (i.e., it directly influences the amount of alcohol consumed in those who have ever drunk alcohol), in a one-sample MR approach within 261,991 adults of European ancestry (with 14,549 cases) (23). This is a useful approach as there is unlikely to be bias due to horizontal pleiotropy. Therefore, further exploration, using two-sample MR and exploring underlying continuous traits may strengthen the conclusion of no effect, if results were consistent. The second study also used a one-sample MR approach and a genetic risk score for alcohol within UK Biobank (UKBB) (N=408,540 with 33,656 cases) and found a potential causal effect of higher alcohol intake on T2D risk, with the strongest effects found in heavier drinkers in analyses stratified by alcohol intake (24). Our study extends this work by including continuous traits related to diabetes and examining this within a two-sample MR approach as well.

The aim of this study was to use MR to explore the effects of lifetime smoking and alcohol consumption (drinks per week) on T2D risk and underlying glycaemic traits. This adds to previous studies by exploring both T2D and related continuous traits, exploring effects of both smoking and alcohol behaviours in the same study, undertaking more sensitivity analyses to test genetic instrument validity, and the influence of unbalanced horizontal pleiotropy on our results, and comparing results from our main two-sample MR with those from one-sample MR, where this was possible, and data was available. In this study we used lifetime smoking as one exposure as this can be applied to non-smokers (unlike smoking heaviness) too and allows for a richer phenotype incorporating a range of smoking behaviours. We used alcohol consumption as our other exposure (i.e., average number of drinks participants reported consuming each week across all types of alcohol) to capture drinking over the whole distribution.

## Methods

### Exposure GWAS and selection of genetic instruments

We used the largest GWAS of lifetime smoking index (LSI) (25) and alcohol consumption (drinks per week) (26), avoiding sample overlap with the outcome GWAS, which can bias results. Both GWAS only included participants of European ancestry and with complete genotype and phenotypic data (for relevant smoking and alcohol phenotypes), resulting in 462,690 participants from UKBB in the LSI GWAS and 941,280 participants from GSCAN (GWAS and Sequencing Consortium of Alcohol and Nicotine) in the drinks per week GWAS. GWAS adjusted for principal components to further control for population substructure.

SNPs (and associations with the relevant exposures) were selected if they reached genome- wide statistical significance (p ≤ 5x10-^08^) and were independent (i.e., we excluded SNPs in linkage disequilibrium; r^2^ of 0.001; window of 10,000 kb; European 1000 genomes reference panel). For any palindromic SNPs we tried to infer the positive strand based on allele frequencies, but if this was not possible, these SNPs were excluded. Where a SNP was available for the exposure and not the outcome, we attempted to identify proxy SNPs using LDlink (27) and an LD r^2^ threshold of >0.8. After exclusions and identifying any proxies, we searched for the remaining LSI SNPs in the outcome GWAS (118 SNPs for all outcomes) and the remaining alcohol consumption SNPs in the outcome GWAS (70 SNPs for all outcomes). Details of the exposure GWAS, including derivation of the LSI and drinks per week of alcohol are provided in Supplementary Materials Section 1. To summarise, the LSI reflects a combination of smoking related behaviours including smoking status, duration and heaviness, where never smokers have a score of 0. Drinks per week reflects the average number of drinks/glasses consumed per week by participants.

LSI is in standard deviation (SD) units, therefore, in our MR analyses we explore effects per 1 SD higher LSI. To give context, 1 SD higher LSI value is equivalent to an individual smoking 20 cigarettes per day for 15 years and stopping 17 years ago or smoking 60 cigarettes a day for 13 years and stopping 22 years ago. Natural log-transformed drinks per week were used in the GWAS, therefore, in our MR analyses we explore effects per 1 SD higher log-transformed drinks per week. To give context, in UKBB (the sample used in our one-sample MR) 1 SD was equal to 2.14 of the log transformed drinks per week.

### Outcome GWAS and harmonisation of exposure SNPs

We obtained associations of the exposure SNPs with outcomes from the largest GWAS of T2D (28), fasting glucose (29), fasting insulin (29) and glycated haemoglobin (HbA1c) (29). We only used GWAS data including participants of European ancestry, resulting in 148,726 cases and 965,732 controls from the Million Veteran Program, DIAMANTE and Biobank Japan for T2D. The continuous traits all used data from the Meta-Analyses of Glucose and Insulin-related traits Consortium (MAGIC), with 209,605 participants with data for fasting glucose, 158,550 with data for fasting insulin and 149,289 with data for HbA1c. GWAS summary statistics for the exposure and outcome were harmonised so that the SNP allele- exposure and SNP allele-outcome associations were in the same direction. Details of these GWAS can be found in Supplementary Materials Section 2. To summarise, the T2D GWAS included cases with a diagnosis of T2D and controls without, fasting glucose was measured in mmol/l, fasting insulin in pmol/l in serum and HbA1c as a percentage. Therefore, in our MR analyses results are reported as the odds of T2D and the difference in mean fasting glucose (mmol/l), fasting insulin (pmol/l) and HbA1c (NGSP percent or equivalent) per 1 SD higher LSI or log-transformed drinks per week.

### UK Biobank data for one-sample Mendelian randomisation

We used data from the UKBB, a large population-based prospective health research resource of 503,317 participants (5.5% response of those invited), recruited between 2006 and 2010, aged between 38 and 73 years and from the UK (30). Further details are included in the Supplementary Materials (Section 3 and on the study website (www.ukbiobank.ac.uk)). We were only able to assess the effect of drinks per week on T2D and HbA1c because the LSI genetic instruments were obtained from UKBB and the glucose and insulin measures in UKBB were from non-fasting samples.

#### Drinks per week

The drinks per week phenotype was constructed from responses to questions on the average weekly intake of a range of different alcoholic beverages (defined as number of glasses they had). Where this information was not available, weekly consumption was estimated from measures of average monthly intake (see Supplementary Materials, Section 4 for further details). Data were natural log-transformed due to being right skewed and standardised (1 SD was equal to 2.14 of the log transformed drinks per week).

#### Type 2 diabetes

We derived possible or probable T2D using the Eastwood algorithm (31) (see Supplementary Materials Section 5). In one-sample MR analyses we excluded individuals who had possible or probable type 1 diabetes as per the Eastwood algorithm.

#### HbA1c

Serum HbA1c (mol/mol) was assayed using five Bio-Rad Variant II Turbo analysers, values outside of the reportable range of 15 to 184 mmol/mol, or invalidated for any other reason, were excluded (further information can be found at https://biobank.ctsu.ox.ac.uk/crystal/ukb/docs/serum_hb1ac.pdf). These analysers used high performance liquid chromatography (HPLC) to determine the relative concentration of HbA1c in packed red blood cells, from blood samples (approximately 9ml) collected at recruitment. Many studies examining continuous traits related to T2D exclude participants with a diabetes diagnosis or above thresholds indicative of diabetes. This means that results are not necessarily applicable to the whole population from which the study sample is drawn and can result in selection bias (32). On the other hand, people with a diagnosis of diabetes will have made changes to their lifestyles and/or be on medications that impact these continuous traits and associations with them. Therefore, we conducted analyses with and without excluding those with possible or probable type 1 or type 2 diabetes using the Eastwood algorithm and those who had a HbA1c measure of ≥6.5% (or 48 mmol/mol) at baseline. Data were natural log-transformed due to being right skewed and standardised (1 SD was equal to 0.15 log mmol/mol). As UKBB did not collect fasting samples we have not conducted one-sample MR on fasting glucose and insulin.

#### Genetic data

A total of 488,377 participants had genotyped samples. Pre-imputation quality control, phasing and imputation are described elsewhere (33) and summarised in the Supplementary Materials (Section 6).

### Statistical analysis

We pre-registered the analysis plan for this study on the Open Science Framework in March 2021 (https://osf.io/ygucn). All analyses were conducted in R (34) (version 3.6.2).

#### Two-sample Mendelian randomisation analyses

We conducted two-sample MR analyses using the TwoSampleMR package in R (35).

We used the inverse-variance weighted (IVW) method as our main analysis (36). This fits a linear regression model of the mean SNP-outcome value on mean SNP-exposure value across all SNPs and constrains the intercept of the regression slope to be zero, with the slope providing an unbiased effect estimate under the assumption that there is no horizontal pleiotropy (37). Sensitivity analyses used to explore this assumption were done using MR-Egger (38), weighted median (39), MR Pleiotropy RESidual Sum and Outlier (MR- PRESSO) (40) and Generalised Summary-data-based MR (GSMR) (41) methods.

MR Egger is identical to IVW with the exception that the intercept reflects the best fitted regression model and is not constrained to zero (38). The slope provides a causal estimate controlling for potential unbalanced horizontal pleiotropy. As with IVW, this approach is also subject to the Instrument Strength Independent of Direct Effect (InSIDE) assumption, which may be violated if any pleiotropic effects are all via a single factor that is correlated with the association of the genetic instrument with the exposure (i.e., instrument strength). A non- null intercept from MR Egger is indicative of unbalanced horizontal pleiotropy and we used the p-value for the intercept to assess this.

The weighted median provides an unbiased causal estimate if no more than 50% of the weight of the SNPs used in the genetic instrument are influenced by horizontal pleiotropy (i.e. results may be biased if one single SNP or several SNPs cumulatively contribute 50% of the weight and are horizontally pleiotropic) (39).

MR-PRESSO is used to detect and correct for potential horizontal pleiotropic outliers in the instrument (40). It comprises of three stages to test this. An initial global test assesses whether the total residual sum of squares (RSS) is similar to that expected by chance. Then any outliers are identified by examining the RSS of each SNP. Finally, the extent to which these outliers effect the causal estimate is evaluated by using the distortion test. From this analysis we get an uncorrected and an outlier-corrected causal estimate and we use results from the global and distortion tests to detect horizontal pleiotropy and test for distortion between the estimate before and after correction, respectively. For MR-PRESSO the precision of the p-values is determined by the number of elements to simulate, specified in the model. We used a value of 2,000 for all analyses, except for lifetime smoking on fasting glucose in both main and additional analyses and fasting insulin in the main analyses, where we used 3,000 due to the model being unable to estimate the p-value with the lower value.

The GSMR approach allows estimation of a causal effect including SNPs in the instrument that are correlated, by estimating the LD between SNPs from a reference sample (41). The GSMR model also removes outliers which may be associated with confounding factors by assessing heterogeneity across SNPs, using the heterogeneity in dependent instrument (HEIDI) test, and models the SNP-exposure estimate error which other MR methods do not include.

In addition, we explored between SNP heterogeneity, which might be an indicator of horizontal pleiotropy or violation of other assumptions, using Cochran’s Q, where a p-value <0.05 may indicate the presence of between SNP heterogeneity. We also assessed heterogeneity between SNPs, whilst adjusting for any horizontal pleiotropy for the MR- Egger method, using the Rucker’s Q-test, again with a p-value threshold of <0.05.

The IVW and MR-Egger methods assume that there is no measurement error in the SNP- exposure estimates (39), known as the ‘NO Measurement Error’ (NOME) assumption. The extent of the NOME assumption violation can be quantified using regression dilution I- squared statistics, where a lower value indicates greater violation. An I-squared of less than 0.9, indicates that MR-Egger estimates should be interpreted with caution due to regression dilution and where this is the case, we have conducted simulation extrapolation (SIMEX) corrections as a sensitivity analysis. The SIMEX approach is a bias adjustment method which provides an estimate for the case where NOME had been satisfied. We also estimated the mean F-statistic for each analysis, which indicates instrument strength, where a value under 10 may indicate a weak instrument (39).

Overall, by using these different methods, which make different assumptions, we were able to assess the robustness of evidence for causal effects against violations of the MR assumptions. We were interested in whether there was evidence of causal effects.

However, we have previously shown that causal effect estimates when using exposures related to cigarette smoking may be unreliable (42). Therefore, we considered consistency of evidence (e.g., direction of effect estimate, p-value as a measure of strength of evidence against the null) across analyses to guide our inference regarding whether or not a causal effect may be operating, but did not attempt to directly estimate the magnitude of any such effect (43).

Finally, the GWAS used for the outcomes of fasting glucose and fasting insulin adjust for body mass index (BMI), which can bias our results. Multivariable MR (MVMR) analyses including BMI can help overcome this issue and provide unbiased estimates of the exposure of interest (LSI and drinks per week) on the outcome (44) (see Supplementary Materials Section 7 for further details).

#### One-sample Mendelian randomisation analyses

One-sample MR analyses were conducted using the OneSampleMR and Applied Econometrics with R (AER) packages, respectively, in R (45,46). We generated weighted allele genetic risk scores in UKBB for alcohol consumption using the per-allele regression coefficients from each independent genome-wide significant SNP for each exposure as weights and then summing those weighted values (see Supplementary Materials Section 8). We used two-stage least squares regression with adjustment for age, sex, the first 10 principal components (PCs) (derived from PC analysis of UKBB genotype data, imputed to a reference set combining UK10K haplotype and Haplotype Reference Consortium [HRC] reference panels), assessment centre and genotyping chip. Two genotyping chips were, the UKBB axiom array (which 90% of participants were genotyped with) and the UK BiLEVE array. The latter was used for those in the UK BiLEVE study (47), which was oversampled for smokers, and therefore adjusting for genotyping chip may introduce collider bias. Therefore, we performed analyses with and without adjustment for chip.

### Ethics

All studies that contributed to the exposure and outcome GWAS used in MR analyses had ethics approval and participant consent for their data to be used in genetic analyses. UKBB (data used in one-sample MR analyses) received ethics approval from the UK National Health Service Research Ethics Committee (11/NW/0382).

### Data availability

Access details for the GWAS data used in this study are outlined in Supplementary Table S1. UK Biobank data are available through a procedure described at http://www.ukbiobank.ac.uk/using-the-resource/.

Analysis code is available from the University of Bristol’s Research Data Repository (http://data.bris.ac.uk/data/), at: To be added on publication.

## Results

### Two-sample Mendelian randomisation with lifetime smoking as the exposure

Mean F-statistics for LSI were all 44.27 (Supplementary Table S2). Figure 1, Figure 2 and Supplementary Table S3 provide the results from the main IVW and all sensitivity analyses of the effects of LSI on outcomes. For T2D, the main IVW result suggested a causal effect of higher LSI on T2D risk (OR per 1SD higher LSI=1.42, 95% CI=1.22 to 1.64). Results from weighted median, PRESSO (with and without outlier correction) and the GSMR sensitivity analyses were consistent with this. By contrast MR-Egger and SIMEX adjusted MR-Egger results were in the opposite direction, though with wide confidence intervals (OR per 1SD higher LSI=0.80, 95% CI=0.46 to 1.41 for MR Egger and OR=0.75, 95% CI=0.43 to 1.31 for SIMEX adjusted MR-Egger). There was also evidence of between SNP heterogeneity (Cochran’s Q p-value = 5.42x10^-66^; Rucker’s Q p-value = 1.10x10^-62^) and potential bias due to unbalanced horizontal pleiotropy based on the MR-Egger and SIMEX adjusted MR Egger intercepts (p=0.04 and p=0.02, respectively) and the MR-PRESSO global test (p<0.0003).

**Figure 1.**
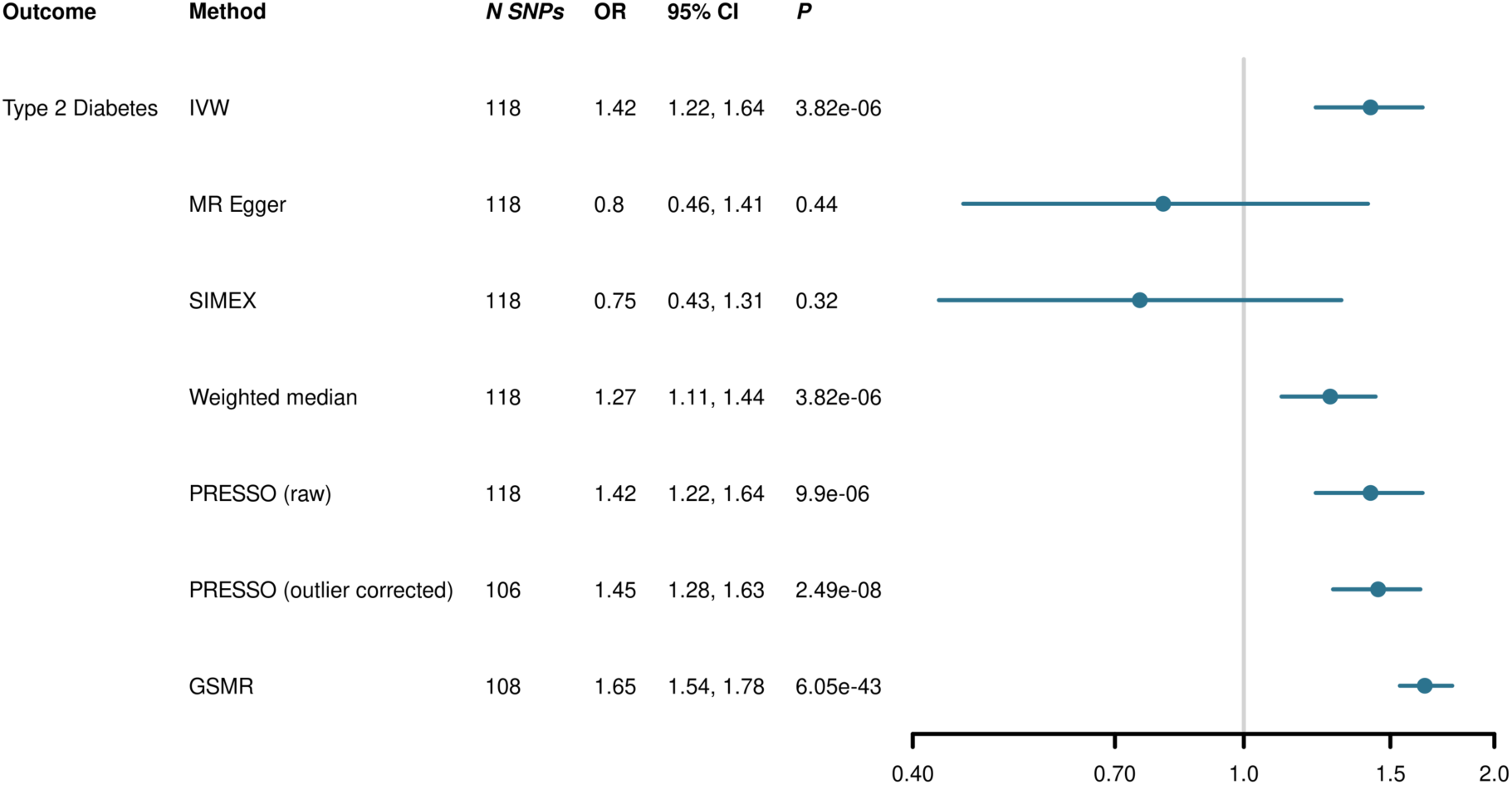
Two-sample Mendelian randomisation results of the potential causal effect of lifetime smoking on type 2 diabetes Results are the odds ratios (OR) of type 2 diabetes per 1 SD higher lifetime smoking index score, with 95% confidence intervals (CI), noting that 1 SD higher LSI value is equivalent to an individual smoking 20 cigarettes per day for 15 years and stopping 17 years ago or smoking 60 cigarettes a day for 13 years and stopping 22 years ago. SNP=single nucleotide polymorphism, IVW=inverse-variance weighted, SIMEX=simulation extrapolation, PRESSO= Pleiotropy RESidual Sum and Outlier, GSMR=Generalised Summary-data-based Mendelian Randomisation

**Figure 2.**
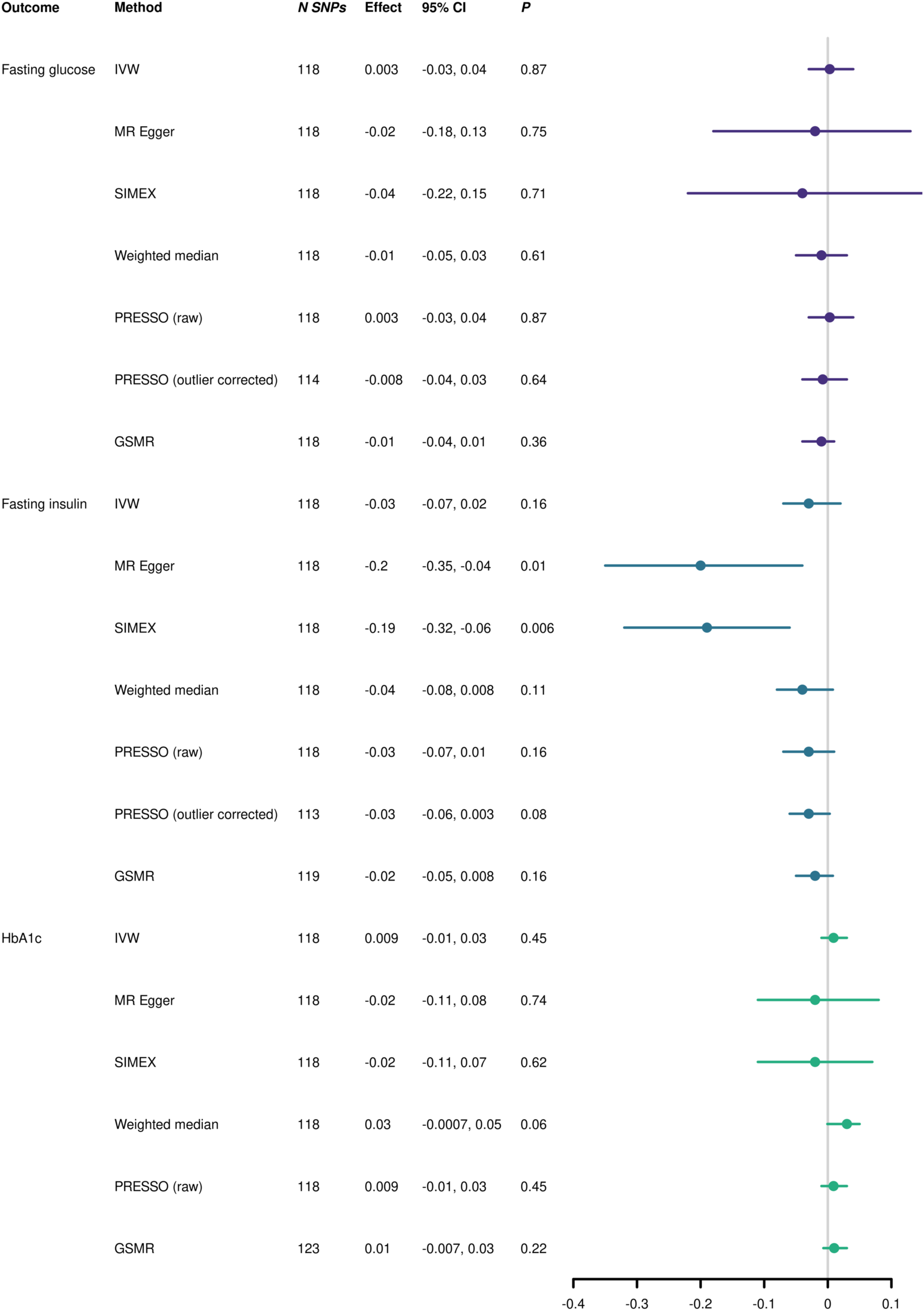
Two-sample Mendelian randomisation results of the potential causal effect of lifetime smoking on glycaemic traits Results are the difference in mean fasting glucose (mmol/l), fasting insulin (pmol/l) and HbA1c (NGSP percent or equivalent) per 1 SD higher lifetime smoking index value, with 95% confidence intervals (CI), noting that 1 SD higher LSI value is equivalent to an individual smoking 20 cigarettes per day for 15 years and stopping 17 years ago or smoking 60 cigarettes a day for 13 years and stopping 22 years ago. SNP=single nucleotide polymorphism, IVW=inverse-variance weighted, SIMEX=simulation extrapolation, PRESSO= Pleiotropy RESidual Sum and Outlier, GSMR=Generalised Summary-data-based Mendelian Randomisation

Our main IVW analyses also suggested that LSI was not causally related to fasting glucose (difference in mean fasting glucose in mmol/l per 1SD higher LSI=0.003, 95% CI=-0.03 to 0.04), fasting insulin (difference in mean fasting insulin in pmol/l per 1SD higher LSI=-0.03, 95% CI=-0.07 to 0.02) or HbA1c (difference in mean HbA1c in NGSP percent or equivalent per 1SD higher LSI=0.009, 95% CI=-0.01 to 0.03). Sensitivity analyses were mostly consistent with these results, with the exception of MR-Egger and SIMEX adjusted MR-Egger, where imprecise estimates suggested that higher LSI might reduce fasting insulin (Figure 2).

Additional MVMR analyses accounting for BMI for fasting glucose and fasting insulin were in line with the main IVW results (Supplementary Table S4).

### Two-sample Mendelian randomisation with drinks per week as the exposure

Mean F-statistics for alcohol consumption were all 31.55 (Supplementary Table S2). Figure 3, Figure 4 and Supplementary Table S3 provide the results from the main IVW and all sensitivity analyses of the effects of alcohol consumption on outcomes. For T2D, the main IVW results suggested that alcohol consumption was not causally related to T2D (OR per 1 SD higher log-transformed drinks per week=1.04, 95% CI=0.40 to 2.65). Some of the sensitivity analyses were consistent with these results, however, MR-Egger, weighted median and GSMR estimates suggested a potential causal effect of higher drinks per week on T2D risk, but confidence intervals were wide (Figure 3). There was also evidence of between SNP heterogeneity (Cochran’s Q p-value = 2.86x10^-89^; Rucker’s Q p-value = 2.00x10^- 70^) and potential bias due to unbalanced horizontal pleiotropy based on the MR Egger intercept (p=0.0005) and the MR-PRESSO global test (p<5x10^-04^).

**Figure 3.**
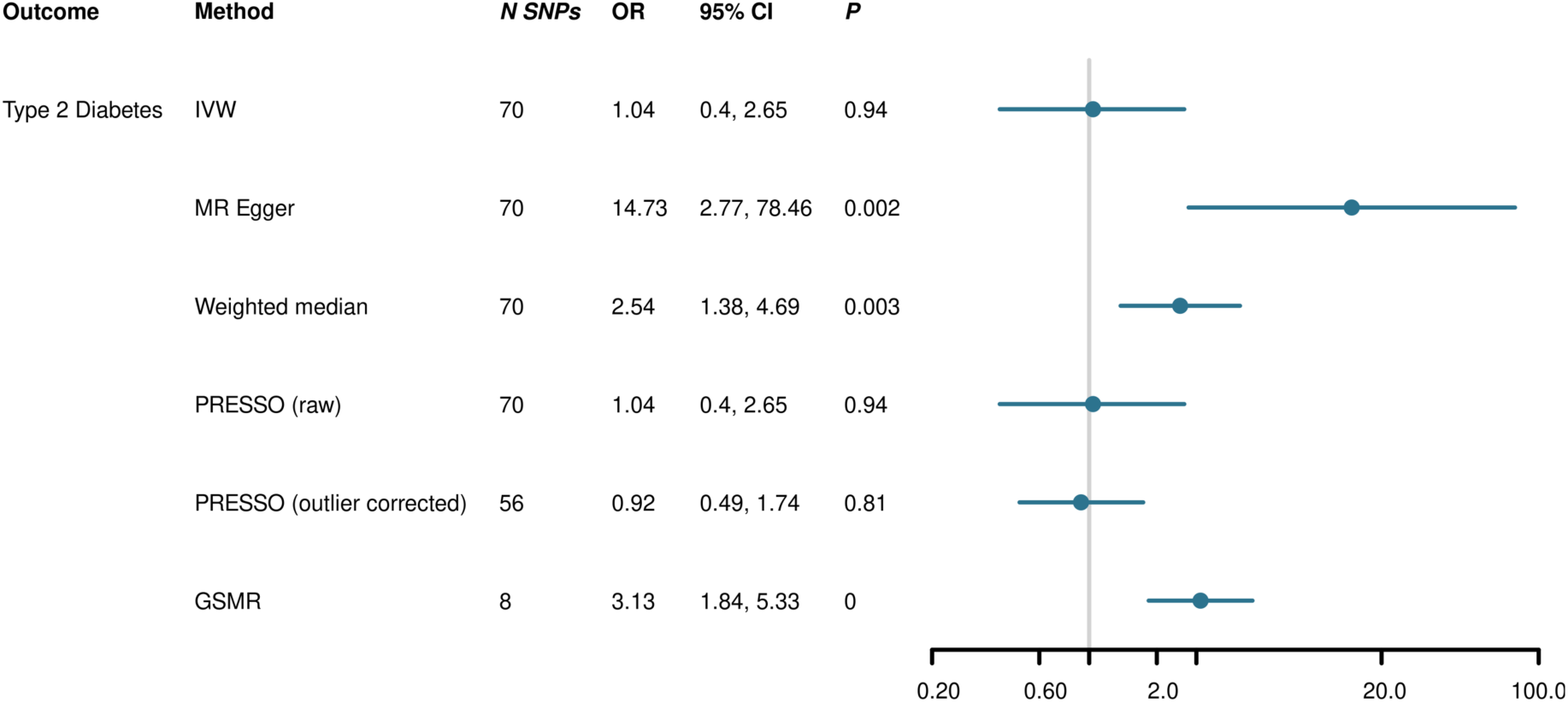
Two-sample Mendelian randomisation results of the potential causal effect of drinks per week on type 2 diabetes Results are the odds ratios (OR) of type 2 diabetes per 1 SD higher log-transformed drinks per week, with 95% confidence intervals (CI). SNP=single nucleotide polymorphism, IVW=inverse-variance weighted, SIMEX=simulation extrapolation, PRESSO= Pleiotropy RESidual Sum and Outlier, GSMR=Generalised Summary-data-based Mendelian Randomisation

**Figure 4.**
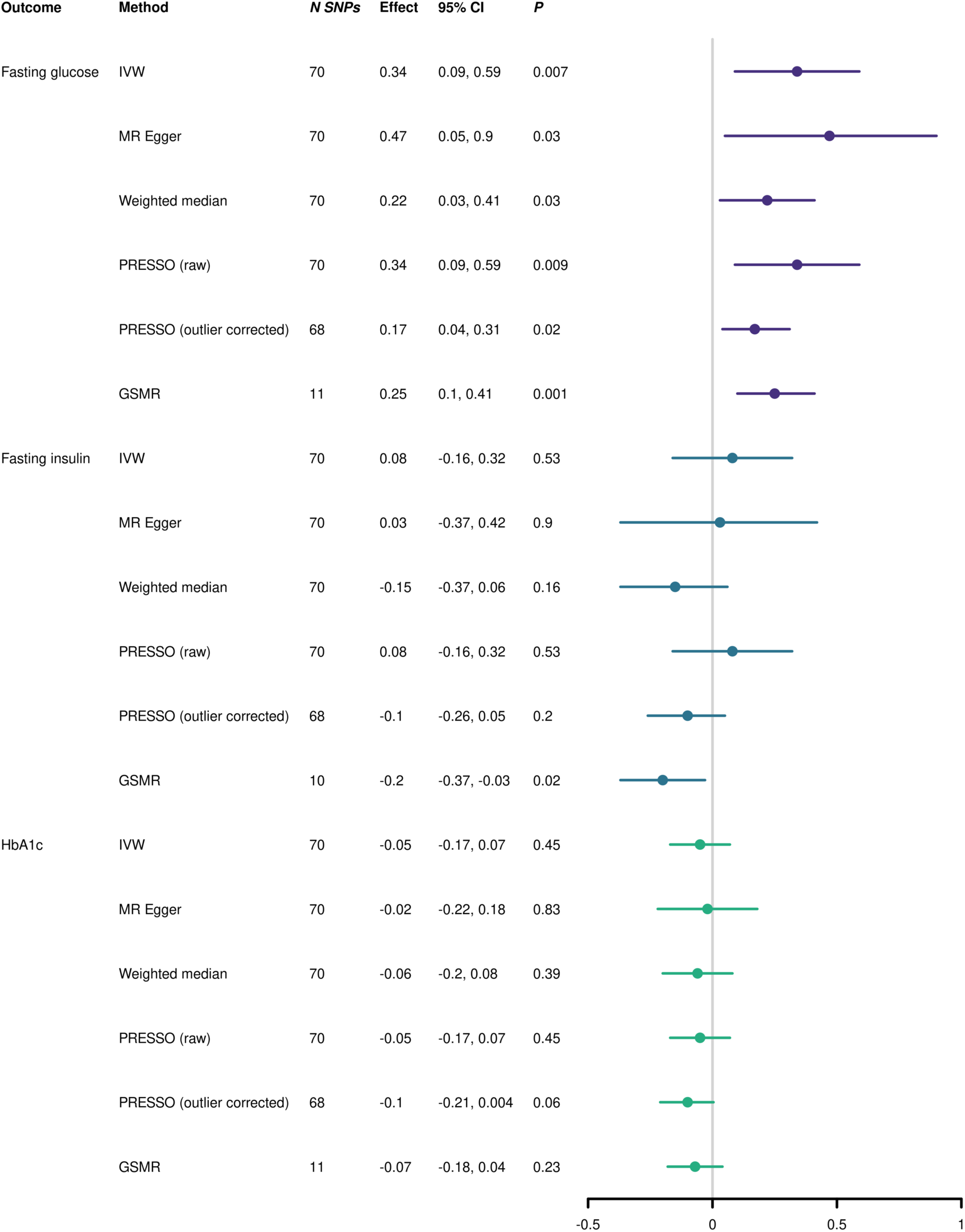
Two-sample Mendelian randomisation results of the potential causal effect of drinks per week on glycaemic traits Results are the difference in mean fasting glucose (mmol/l), fasting insulin (pmol/l) and HbA1c (NGSP percent or equivalent) per 1 SD higher log-transformed drinks per week, with 95% confidence intervals (CI). SNP=single nucleotide polymorphism, IVW=inverse-variance weighted, SIMEX=simulation extrapolation, PRESSO= Pleiotropy RESidual Sum and Outlier, GSMR=Generalised Summary-data-based Mendelian Randomisation

Our main IVW analyses suggested a potential casual effect of higher drinks per week on higher fasting glucose (difference in mean fasting glucose in mmol/l per 1SD higher log-transformed drinks per week=0.34, 95% CI=0.09 to 0.59). This was consistent across all sensitivity analyses. However, there was also evidence of between SNP heterogeneity (Cochran’s Q p-value = 5.18x10^-40^; Rucker’s Q p-value = 7.13x10^-40^) and potential bias due to unbalanced horizontal pleiotropy based on the MR-PRESSO global test (p<5x10^-04^), but not the MR-Egger intercept (p=0.46). Our main IVW analyses suggested that drinks per week was not causally related to fasting insulin (difference in mean fasting insulin in pmol/l per 1SD higher LSI=0.08, 95% CI=-0.16 to 0.32) or HbA1c (difference in mean HbA1c in NGSP percent or equivalent per 1SD higher LSI=-0.05, 95% CI=-0.17 to 0.07). Sensitivity analyses were mostly consistent with these results, with the exception of the GSMR estimate suggesting that higher drinks per week might reduce fasting insulin (Figure 4). The effect from the additional MVMR analyses accounting for BMI for fasting glucose was attenuated and did not provide evidence of a potentially causal effect. For fasting insulin, the MVMR analysis results were in line with the main IVW results (Supplementary Table S4).

#### One-sample Mendelian randomisation for drinks per week on type 2 diabetes and HbA1c

Sample characteristics for those UKBB participants included in the one-sample MR analyses are shown in Supplementary Table S5.

Table 1 provides the results from the one-sample MR analyses. Results suggested a causal effect of higher drinks per week on T2D risk (OR per 1 SD higher log-transformed drinks per week=1.71, 95% CI: 1.24 to 2.36) and on lower HbA1c levels (but only when we excluded participants with a possible or probable diabetes diagnosis or HbA1c≥6.5%) (difference in mean SD of log transformed HbA1c (mol/mol) per 1 SD higher log-transformed drinks per week=-0.07, 95% CI: -0.11 to -0.02). Adjusting for chip did not impact our results.

**Table 1.**
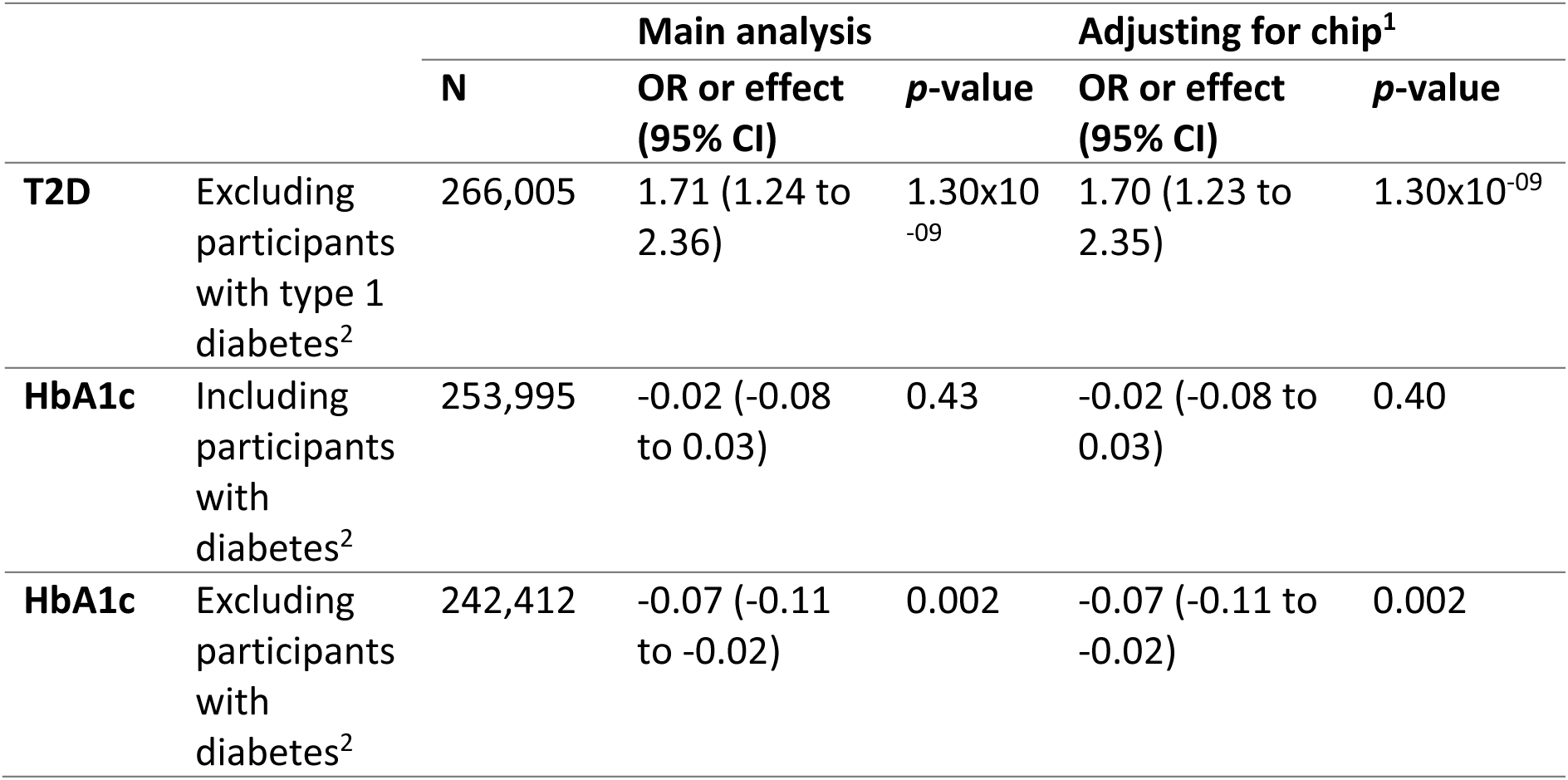
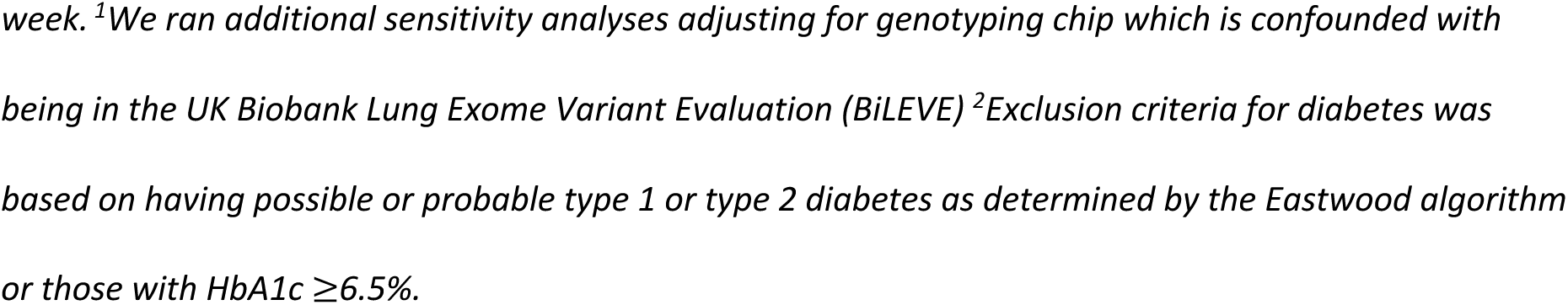
One-sample Mendelian randomisation results.

## Discussion

We found evidence of a possible causal effect of higher LSI values on risk of T2D in our main IVW analysis, but this was not consistent across sensitivity analyses and may be biased due to unbalanced horizontal pleiotropy. Furthermore, the lack of evidence of an effect on the underlying glycaemic traits suggests that we cannot confidently interpret this as a causal effect on T2D risk. We found little evidence of a possible causal effect of higher drinks per week on T2D risk, with no effect found for our main IVW analyses in the two-sample MR and inconsistent evidence in our sensitivity analyses. However, the lack of a consistent finding means that we would not interpret this as being evidence of a causal effect. We did observe a possible causal effect of drinks per week on fasting glucose, which was consistent across sensitivity analyses, but this was not supported by analyses with the other underlying glycaemic traits and when accounting for BMI this effect was no longer observed.

Additionally, results from our one-sample MR analyses (only examining drinks per week on T2D and HbA1c) suggest that there may be a causal effect of more drinks per week on T2D risk and lower HbA1c levels. These mixed results do not provide strong evidence of causal effects but do suggest that LSI and drinks per week may have some effect on some of these outcomes; however, the explanations for these effects may be complicated, for example, acting through pleiotropic pathways.

Previous studies have largely shown associations using multivariable regression, and potential causal effects using MR, between smoking and T2D risk (10,12,21,22,48) and between higher alcohol consumption and T2D risk (17,18,23,24). However, we note that a previous MR study using a functional variant for alcohol metabolism did not find an effect on T2D (23), in line with our results, suggesting it is unlikely there is an effect of alcohol on T2D. In addition, our finding with decreased HbA1c in the one-sample MR analyses is in the opposite direction to what we might expect given that higher levels of HbA1c are observed in those with T2D. However, this may be due to the exclusion of individuals with a diagnosis of diabetes, and we do not find an effect in our two-sample MR, so results should be interpreted with caution. Our results suggest that future studies examining risk factors for T2D should triangulate results across different analyses and consider underlying glycaemic traits as outcomes in order to arrive at the correct conclusions.

### Limitations

The key strengths of this study and how this study adds to previous literature is described in the introduction and above. In terms of limitations, we were unable to explore a non-linear effect as recent evidence suggests that current methods for doing so in MR are potentially biased (49). Some observational studies have suggested a J-shaped association between alcohol and coronary heart disease, which T2D is a risk factor for (14–16,19). However, that J-shaped association, even if causal, suggests a linear association across most of the distribution. Our one-sample MR analyses in UKBB may be subject to selection bias (50,51). Future studies replicating these analyses in other samples would be useful to examine whether selection bias in UKBB may be an issue here, however our two-sample MR analyses in part overcome this for alcohol but not lifetime smoking where the GWAS was conducted in UK Biobank as well. Our analyses were conducted in samples of European ancestry, due to the data available, so results are not generalisable beyond this group. Finally, it is possible that measurement error in the exposure or outcome could bias our results. This is more likely to be the case for the exposures, which could be subjectively influenced, for example misreporting of cigarettes smoked per day, time since cessation or duration of smoking and number of drinks consumed per week. In particular, in UKBB drinks per week is measured based on number of glasses of alcohol consumed and this is similar to the definitions used in the GWAS. This doesn’t account for units and glass/drink size can vary, so there is likely variation in how this was reported.

## Conclusion

In summary, we found limited evidence of a possible causal effect of higher lifetime smoking index score and drinks per week score on T2D risk. We found further evidence of a possible causal effect of higher drinks per week on higher fasting glucose. However, overall results were not consistent across analyses and some results may be biased by horizontal pleiotropy. Therefore, we do not find strong evidence of smoking and alcohol influencing risk of T2D. Future research should include triangulation approaches and glycaemic traits to allow for a more in depth understanding of the causal influence of risk factors on T2D.

## Data Availability

Access details for the GWAS data used in this study are outlined in Supplementary Table S1. UK Biobank data are available through a procedure described at http://www.ukbiobank.ac.uk/using-the-resource/.
Analysis code is available from the University of Bristol's Research Data Repository (http://data.bris.ac.uk/data/), at: To be added on publication.

## Acknowledgements

This research has been conducted using data from UKBB (project ID: 9142), a major biomedical database (http://www.ukbiobank.ac.uk/). We would like to thank the research participants and employees of 23andMe, inc. for making this work possible. For the purpose of open access, the author(s) has applied a Creative Commons Attribution (CC BY) licence to any Author Accepted Manuscript version arising from this submission. The type 2 diabetes GWAS included data from the Million Veteran Program (MVP), Office of Research and Development, Veterans Health Administration, and was supported by the Veterans Administration (VA). The authors thank MVP staff, researchers, and volunteers, who have contributed to MVP, and especially participants who previously served their country in the military and now generously agreed to enroll in the study. (See https://www.research.va.gov/mvp/ for more details). The MVP GWAS data was provided through dbGaP under accession number phs001672.

## Author contributions

Conceptualization: ASA, MRM, DAL; Methodology: ZER, HMS, MRM, DAL; Data Curation: ZER, RCR; Formal Analysis: ZER; Investigation: ZER; Resources: ZER, MRM; Writing—Original Draft: ZER; Writing—Review and Editing: ZER, HMS, RCR, ASA, DAL, MRM; Supervision: DAL, MRM; Project Administration: ZER; Funding Acquisition: DAL, MRM.

