## Supplementary Materials for "Do smoking and alcohol behaviours influence risk of type 2 diabetes? A Mendelian randomisation study"

*1. Exposure GWAS details*

*Lifetime smoking index (LSI):* A GWAS conducted in UK Biobank (UKBB) (N= 462,690) (Wootton et al., 2019). The LSI captures information on smoking status, duration and heaviness, derived by combining this information with a simulated half-life constant of 18 (capturing the decreasing effect of smoking at a given time on health outcomes), where individuals who had never smoked had a value of 0. Genome wide associations with LSI in standard deviation (SD) units were estimated and used in our MR analyses.

*Drinks per week:* A GWAS conducted in GSCAN (GWAS and Sequencing Consortium of Alcohol and Nicotine, N=941,280) (Liu et al., 2019). Participants responded to questions regarding the number of alcoholic beverages consumed in the past week and average number per week in the past year. Drinks were defined differently across studies but generally do not refer to units but instead drinks/glasses. Average number of drinks per week was calculated and data were log-transformed. This GWAS used data from several cohorts. We used all 99 SNPs (where available) that were genome-wide significant in the full sample in the genetic risk scores for both our two-sample and one-sample MR. However, to avoid sample overlap in our MR analyses between the exposure and outcome GWAS used, when creating these genetic risk scores, we used weights of these SNPs from the GWAS for the 23andMe cohort only for two-sample MR (N=403,931) and weights from the GWAS for all cohorts except UKBB for one-sample MR (N=630,154).

*2. Outcome GWAS details*

*T2D:* We used a meta-analysis of GWAS that used data from several studies including the Million Veteran Program (MVP; discovery cohort), DIAMANTE and Biobank Japan (N=148,726 cases and 965,732 controls of European ancestry) (Vujkovic et al., 2020). Cases were those with a diagnosis of T2D.

*Continuous traits:* We used data from the Meta-Analyses of Glucose and Insulin-related traits Consortium (MAGIC) from the European ancestry subset. We used the paper by Chen et al (Chen et al., 2021) for fasting glucose, measured in mmol/l (N=209,605), fasting insulin, measured in pmol/l in serum, (N=158,550 of European ancestry), and HbA1c, measured as a percentage (N=149,289 of European ancestry). In cohorts where fasting glucose was measured in whole blood, values were equivalised to plasma level by multiplying by 1.13. HbA1C was reported as a National Glycohemoglobin Standardization Program (NGSP) percent or standardised in a similar manner, for example with the Direct Control and Complications Trial (DCCT) reference method, where this was required. Participants for all outcomes were excluded if they had a physician diagnosis of diabetes, were on diabetes treatment (oral or insulin), or had fasting glucose $\geq$7mmol/l, or if this was unavailable then excluded if 2hr glucose $\geq$11.1mmol/l or HbA1c $\geq$6.5%.

*3. UK Biobank description*

In our one-sample Mendelian randomisation (MR) analyses we used data from the UK Biobank. Baseline assessments took place at 22 assessment centres in the UK. Data collected includes sociodemographic, lifestyle and physical and mental health measures. Data have been collected via questionnaires, computer assisted interviews, clinic visits and data linkage and further data collection is ongoing.

In our analyses we excluded any participants who withdrew their consent for their data to be used by using the latest withdrawal lists provided for the project data we have access to (UKBB project number: 9142). We also restricted analyses to individuals of “White British” ancestry and removed related individuals.

*4. Drinks per week phenotype*

The construction of this variable was the same as that used for the drinks per week GWAS in UKBB (Liu et al., 2019). Participants were asked several questions related to alcohol consumption in the touchscreen questionnaires completed at the assessment centres. Data was only collected from participants who indicated that they drank alcohol more often than once or twice a week. Participants were asked how many glasses/pints/measures of red wine, champagne and white wine, beer and cider, spirits, fortified wine and other alcoholic beverages they consumed in an average week in separate questions. Participants who responded with ‘do not know’ or ‘prefer not to answer’ were assigned a value of 0. The responses given for beer or cider were multiplied by 1.3 and the responses given for other were multiplied by 1.5, as per the Liu et al drinks per week GWAS (Liu et al., 2019) using this variable. These were adjusted based on the average size of a pint and average UK portion sizes and percentage alcohol by volume for the other beverages. Where data was not available for drinks per week, but was available for drinks per month, this value divided by the average number of days in a month (30.42) times days in the week (7). If this value was greater than 168, this was set to missing.

*5. Other UK Biobank data used for excluding individuals in one-sample MR*

Details on the Eastwood algorithm can be found in the paper describing it (Eastwood et al., 2016). However, to summarise, the algorithm is used to identify diabetes status using a combination of self-report, primary and secondary care data measures within UK Biobank. We used this to identify ‘possible’ and ‘probable’ cases of type 1 and type 2 diabetes, where probable cases have stronger evidence of this being the case from primary care data and possible cases had weaker evidence.

*6. Genetic data from UK Biobank*

Multiallelic SNPs and those with a minor allele frequency (MAF) ≤1% were removed. Genotype data phasing was performed using a modified version of the SHAPEIT2 algorithm. SNPs were imputed to the Haplotype Reference Consortium (HRC) reference panel, using IMPUTE2 algorithms. Graded filtering of SNPs was used, with different imputation qualities for different MAF ranges (Info>0.3 for MAF>3%, Info>0.6 for MAF 1-3%, Info>0.8 for MAF 0.5-1% and Info>0.9 for MAF 0.1-0.5%), where info and MAF scores were recalculated on an in-house derived ‘European’ subset. Individuals with sex-mismatch or sex-chromosome aneuploidy were excluded (N=814). In-house quality control filtering of data is described in a published protocol (Mitchell et al., 2019).

We restricted our sample to individuals of “White British” ancestry or who have very similar ancestral backgrounds according to PC analysis (N=409,703) (Bycroft et al., 2018). Kinship coefficients were estimated with the KING toolset (Manichaikul et al., 2010) and 107,162 pairs of related individuals were identified. An in-house algorithm was applied to this list and removed those individuals related to the greatest number of other individuals until no related pairs remain (N=79,448). Additionally, 2 individuals were removed due to them relating to a very large number (>200) of individuals.

*7. Multivariable Mendelian randomisation to account for BMI*

Where fasting glucose and fasting insulin were the outcomes we also conducted multivariable MR (MVMR) analyses to provide unbiased estimates for the effects of LSI and drinks per week on these outcomes, accounting for BMI (Gilbody et al., 2022).

For BMI, we used data from a GWAS conducted in UK Biobank, from the IEU Open GWAS project (ukb-b-2303; N=456,426) (Elsworth et al., 2020).

We used the *MVMR* package in R (Sanderson et al., 2021) to conduct IVW MVMR analyses.

*8. Genetic risk score construction for one-sample Mendelian randomisation analyses*

Genetic risk scores were constructed using Plink (version 2) (Chang et al., 2015). SNPs from the drinks per week GWAS and UKBB were harmonised and clumped using the European subsample of the 1000 genomes project, with r^2^ <0.25 and a window of 500 kb. Risk scores were created for each participant by multiplying the number of effect alleles by the effect estimate of the SNP, then summing across all SNPs. Genetic risk scores were z-standardised.

Supplementary Table S1. Data availability for the genome-wide association studies used

| **Phenotype** | **Data access information** |
| --- | --- |
| Lifetime smoking index | Available from: <https://data.bris.ac.uk/data/dataset/10i96zb8gm0j81yz0q6ztei23d> |
| Drinks per week | GWAS data excluding 23andMe and excluding UK Biobank can be accessed here: <https://conservancy.umn.edu/handle/11299/201564>  Full GWAS summary statistics for the 23andMe discovery data set will be made available through 23andMe to qualified researchers under an agreement with 23andMe that protects the privacy of the 23andMe participants. Please visit <https://research.23andme.com/collaborate/#dataset-access/> for more information and to apply to access the data. |
| Diabetes (type 2) | GWAS data are available through dbGaP under accession number phs001672.v3.p1 (Veterans Administration Million Veteran Program Summary Results from Omics Studies). Data access must be requested via dbGaP (<https://dbgap.ncbi.nlm.nih.gov/aa/wga.cgi?page=login>). |
| Fasting glucose, fasting insulin and HbA1c | Available from: <https://magicinvestigators.org/downloads/> |

Supplementary Table S2. Mean F statistic and weighted and unweighted I-squared results for Mendelian randomisation analyses.

| Exposure | Outcome | Mean F Statistic | Unweighted I-squared | Weighted I-squared |
| --- | --- | --- | --- | --- |
| Lifetime smoking | Diabetes | 44.27 | 0.65 | 0.63 |
|  | Fasting glucose | 44.27 | 0.65 | 0.46 |
|  | Fasting insulin | 44.27 | 0.65 | 0.46 |
|  | HbA1c | 44.27 | 0.65 | 0.50 |
| Drinks per week | Diabetes | 31.55 | 0.92 | 0.88 |
|  | Fasting glucose | 31.55 | 0.92 | 0.90 |
|  | Fasting insulin | 31.55 | 0.92 | 0.90 |
|  | HbA1c | 31.55 | 0.92 | 0.90 |

*The mean F-statistic is indicative of instrument strength. A value under 10 indicates that there may be a weak instrument. I-squared values greater than 0.9 indicate minimal bias in the MR-Egger estimate. For values less than 0.9 (if both weighted and unweighted were below 0.9) we ran simulation extrapolation (SIMEX) corrections and report these in addition to our MR-Egger results. We therefore ran simulation extrapolation (SIMEX) corrections for all the lifetime smoking on glycaemic trait analyses.*

Supplementary Table S3. Two-sample Mendelian randomisation results with lifetime smoking and drinks per week as the exposures and diabetes, fasting glucose, fasting insulin and HbA1c as the outcomes.

| Exposure | Outcome | Method | N SNPs | OR or effect estimate^1^ (95% CI) | *p-*value | Heterogeneity test *p*-value^2^ | Directional pleiotropy intercept (95% CI; *p*-value) | Global and distortion tests for MR PRESSO |
| --- | --- | --- | --- | --- | --- | --- | --- | --- |
| Lifetime smoking | Diabetes | IVW | 118 | 1.42 (1.22 to 1.64) | 3.82x10^-06^ | 5.42x10^-66^ |  |  |
|  |  | MR Egger | 118 | 0.80 (0.46 to 1.41) | 0.44 | 1.10x10^-62^ | 0.009 (0.0004 to 0.02; 0.04) |  |
|  |  | SIMEX adjusted MR-Egger^3^ | 118 | 0.75 (0.43 to 1.31) | 0.32 | - | 0.01 (0.002 to 0.02; 0.02) |  |
|  |  | Weighted median | 118 | 1.27 (1.11 to 1.44) | 3.82x10^-06^ |  |  |  |
|  |  | PRESSO - raw | 118 | 1.42 (1.22 to 1.64) | 9.90x10^-06^ |  |  | Global = 611.32; *p*<0.0003  Distortion = -5.85; *p*=0.73 |
|  |  | PRESSO – outlier corrected | 106 (12 outliers) | 1.45 (1.28 to 1.63) | 2.49x10^-08^ |  |  |  |
|  |  | GSMR | 108 | 1.65 (1.54 to 1.78) | 6.05x10^-43^ |  |  |  |
|  | Fasting glucose | IVW | 118 | 0.003 (-0.03 to 0.04) | 0.87 | 4.80x10^-13^ |  |  |
|  |  | MR Egger | 118 | -0.02 (-0.18 to 0.13) | 0.75 | 3.45x10^-13^ | 0.0004 (-0.002 to 0.003; 0.71) |  |
|  |  | SIMEX adjusted MR-Egger^3^ | 118 | -0.04 (-0.22, 0.15) | 0.71 | - | 0.0005 (-0.002 to 0.003; 0.74) |  |
|  |  | Weighted median | 118 | -0.01 (-0.05 to 0.03) | 0.61 |  |  |  |
|  |  | PRESSO - raw | 118 | 0.003 (-0.03 to 0.04) | 0.87 |  |  | Global = 266.20; *p*<0.0003  Distortion = 137.45; *p*=0.22 |
|  |  | PRESSO – outlier corrected | 114 (4 outliers) | -0.008 (-0.04 to 0.03) | 0.64 |  |  |  |
|  |  | GSMR | 118 | -0.01 (-0.04 to 0.01) | 0.36 |  |  |  |
|  | Fasting insulin | IVW | 118 | -0.03 (-0.07 to 0.02) | 0.16 | 6.64x10^-08^ |  |  |
|  |  | MR Egger | 118 | -0.20 (-0.35 to -0.04) | 0.01 | 3.96x10^-07^ | 0.003 (0.0003 to 0.005; 0.03) |  |
|  |  | SIMEX adjusted MR-Egger^3^ | 118 | -0.19 (-0.32, -0.06) | 0.006 | - | 0.003 (0.0005 to 0.005; 0.02) |  |
|  |  | Weighted median | 118 | -0.04 (-0.08 to 0.008) | 0.11 |  |  |  |
|  |  | PRESSO - raw | 118 | -0.03 (-0.07 to 0.01) | 0.16 |  |  | Global = 220.03; *p*<0.0003  Distortion = 2.51; 0.96 |
|  |  | PRESSO – outlier corrected | 113 (5 outliers) | -0.03 (-0.06 to 0.003) | 0.08 |  |  |  |
|  |  | GSMR | 119 | -0.02 (-0.05 to 0.008) | 0.16 |  |  |  |
|  | HbA1c | IVW | 118 | 0.009 (-0.01 to 0.03) | 0.45 | 0.0002 |  |  |
|  |  | MR-Egger | 118 | -0.02 (-0.11 to 0.08) | 0.74 | 0.0002 | 0.0004 (-0.001 to 0.002; 0.60) |  |
|  |  | SIMEX adjusted MR-Egger^3^ | 118 | -0.02 (-0.11 to 0.07) | 0.62 | - | 3.81x10^-04^ (-0.001 to 0.002; 0.59) |  |
|  |  | Weighted median | 118 | 0.03 (-0.0007 to 0.05) | 0.06 |  |  |  |
|  |  | PRESSO - raw | 118 | 0.009 (-0.01 to 0.03) | 0.45 |  |  | Global = 180.79; *p*<0.0003  Distortion = NA |
|  |  | PRESSO – outlier corrected^4^ | NA | NA | NA |  |  |  |
|  |  | GSMR | 123 | 0.01 (-0.007 to 0.03) | 0.22 |  |  |  |
| Drinks per week | Diabetes | IVW | 70 | 1.04 (0.40 to 2.65) | 0.94 | 2.86x10^-89^ |  |  |
|  |  | MR-Egger | 70 | 14.73 (2.77 to 78.46) | 0.002 | 2.00x10^-70^ | -0.01 (-0.02 to -0.006; 0.0005) |  |
|  |  | Weighted median | 70 | 2.54 (1.38 to 4.69) | 0.003 |  |  |  |
|  |  | PRESSO - raw | 70 | 1.04 (0.40 to 2.65) | 0.94 |  |  | Global = 652.61; *p*<5x10^-04^  Distortion = 145.101; *p*=0.15 |
|  |  | PRESSO – outlier corrected | 56 (14 outliers) | 0.92 (0.49 to 1.74) | 0.81 |  |  |  |
|  |  | GSMR^5^ | 8 | 3.13 (1.84 to 5.33) | 2.56x10^-05^ |  |  |  |
|  | Fasting glucose | IVW | 70 | 0.34 (0.09 to 0.59) | 0.007 | 5.18x10^-40^ |  |  |
|  |  | MR Egger | 70 | 0.47 (0.05 to 0.90) | 0.03 | 7.13x10^-40^ | -0.0007 (-0.003 to 0.001; 0.46) |  |
|  |  | Weighted median | 70 | 0.22 (0.03 to 0.41) | 0.03 |  |  |  |
|  |  | PRESSO - raw | 70 | 0.34 (0.09 to 0.59) | 0.009 |  |  | Global = 378.69; *p*<5x10^-04^  Distortion = 96.31; *p*=0.002 |
|  |  | PRESSO – outlier corrected | 68 (2 outliers) | 0.17 (0.04 to 0.31) | 0.02 |  |  |  |
|  |  | GSMR | 11 | 0.25 (0.10 to 0.41) | 0.001 |  |  |  |
|  | Fasting insulin | IVW | 70 | 0.08 (-0.16 to 0.32) | 0.53 | 9.53x10^-25^ |  |  |
|  |  | MR Egger | 70 | 0.03 (-0.37 to 0.42) | 0.90 | 5.57x10^-25^ | 0.0003 (-0.002 to 0.002; 0.75) |  |
|  |  | Weighted median | 70 | -0.15 (-0.37 to 0.06) | 0.16 |  |  |  |
|  |  | PRESSO - raw | 70 | 0.08 (-0.16 to 0.32) | 0.53 |  |  | Global = 284.66; *p*<5x10^-04^  Distortion = 175.61; *p*=0.26 |
|  |  | PRESSO – outlier corrected | 68 (2 outliers) | -0.10 (-0.26 to 0.05) | 0.20 |  |  |  |
|  |  | GSMR | 10 | -0.20 (-0.37 to -0.03) | 0.02 |  |  |  |
|  | HbA1c | IVW | 70 | -0.05 (-0.17 to 0.07) | 0.45 | 8.64x10^-08^ |  |  |
|  |  | MR-Egger | 70 | -0.02 (-0.22 to 0.18) | 0.83 | 6.08x10^-08^ | -0.0001 (-0.001 to 0.0008, 0.77) |  |
|  |  | Weighted median | 70 | -0.06 (-0.20 to 0.08) | 0.39 |  |  |  |
|  |  | PRESSO - raw | 70 | -0.05 (-0.17 to 0.07) | 0.45 |  |  | Global = 152.84; *p*<5x10^-04^  Distortion = 55.58; *p*=0.44 |
|  |  | PRESSO – outlier corrected | 68 (2 outliers) | -0.10 (-0.21 to 0.004) | 0.06 |  |  |  |
|  |  | GSMR | 11 | -0.07 (-0.18 to 0.04) | 0.23 |  |  |  |

*^1^Results are the odds ratios (OR) of type 2 diabetes per 1 SD higher lifetime smoking index or per 1 SD higher log-transformed drinks per week, with 95% confidence intervals (CI) or the difference in mean fasting glucose (mmol/l), fasting insulin (pmol/l) and HbA1c (NGSP percent or equivalent) per 1 SD higher lifetime smoking index or per 1 SD higher log-transformed drinks per week, with 95% confidence intervals (CI). To give context, 1 SD higher LSI value is equivalent to an individual smoking 20 cigarettes per day for 15 years and stopping 17 years ago or smoking 60 cigarettes a day for 13 years and stopping 22 years ago.*

*^2^Heterogeneity between SNPs was assessed using the Cochran’s test for the IVW method and Rucker’s Q-test for the MR-Egger method.*

*^3^The I-squared value was <0.9, indicating a potential violation of the NO Measurement Error (NOME) assumption. We ran SIMEX corrected MR-Egger and present these results in addition to the main MR-Egger results to allow for estimates where NOME is not violated.*

*^4^Where NA is presented for the MR-PRESSO outlier-corrected method this indicates that the global test did not detect any pleiotropy and therefore the outlier-corrected method was not required.*

*^5^Note that this analysis included less than the recommended 10 SNPs for GSMR*

*SNP=Single nucleotide polymorphism, OR=Odds ratio, CI=Confidence interval, MR=Mendelian randomisation, PRESSO=Pleiotropy RESidual Sum and Outlier, IVW=Inverse-variance weighted, GSMR=Generalised Summary-data-based MR.*

Supplementary Table S4. Multivariable Mendelian randomisation results for analyses accounting for body mass index.

| Exposure | Outcome | Effect estimate^1^ (95% CI) | *p-*value |
| --- | --- | --- | --- |
| Lifetime smoking | Fasting glucose | 0.001 (-0.04 to 0.04) | 0.96 |
| BMI | Fasting glucose | -0.005 (-0.02 to 0.01) | 0.54 |
| Lifetime smoking | Fasting insulin | -0.007 (-0.05 to 0.04) | 0.75 |
| BMI | Fasting insulin | -0.03 (-0.04 to -0.007) | 0.008 |
| Drinks per week | Fasting glucose | 0.12 (-0.06 to 0.30) | 0.18 |
| BMI | Fasting glucose | 0.001 (-0.02 to 0.02) | 0.86 |
| Drinks per week | Fasting insulin | -0.09 (-0.27 to 0.10) | 0.35 |
| BMI | Fasting insulin | -0.02 (-0.04 to -0.007) | 0.006 |

*^1^Results are the difference in mean fasting glucose (mmol/l) and fasting insulin (pmol/l) per 1 SD higher lifetime smoking index or per 1 SD higher log-transformed drinks per week, with 95% confidence intervals (CI). To give context, 1 SD higher LSI value is equivalent to an individual smoking 20 cigarettes per day for 15 years and stopping 17 years ago or smoking 60 cigarettes a day for 13 years and stopping 22 years ago.*

*CI=Confidence interval, BMI=body mass index*

Supplementary Table S5. Summary statistics for UK Biobank sample with data available.

| Phenotype | N | Mean (SD) or percentage (%) |
| --- | --- | --- |
| Age (years) | 336,984 | 56.87 (8.00) |
| Sex (females) | 336,984 | 53.79% |
| Probable or possible type 1 diabetes | 336,984 | 0.33% |
| Probable or possible type 2 diabetes | 336,984 | 4.29% |
| HbA1c | 321,063 | 35.96 (6.50) |
| Drinks per week | 266,829 | 11.36 (10.87) |
| Genotype chip confounded with being in BiLEVE | 336,984 | 9.24% |

*SD=standard deviation, HbA1c=* *glycated haemoglobin, BiLEVE= UK Biobank Lung Exome Variant Evaluation*

Chen, J., Spracklen, C.N., Marenne, G., Varshney, A., Corbin, L.J., Luan, J., Willems, S.M., Wu, Y., Zhang, X., Horikoshi, M., Boutin, T.S., Mägi, R., Waage, J., Li-Gao, R., Chan, K.H.K., Yao, J., Anasanti, M.D., Chu, A.Y., Claringbould, A., Heikkinen, J., Hong, J., Hottenga, J.J., Huo, S., Kaakinen, M.A., Louie, T., März, W., Moreno-Macias, H., Ndungu, A., Nelson, S.C., Nolte, I.M., North, K.E., Raulerson, C.K., Ray, D., Rohde, R., Rybin, D., Schurmann, C., Sim, X., Southam, L., Stewart, I.D., Wang, C.A., Wang, Y., Wu, P., Zhang, W., Ahluwalia, T.S., Appel, E.V.R., Bielak, L.F., Brody, J.A., Burtt, N.P., Cabrera, C.P., Cade, B.E., Chai, J.F., Chai, X., Chang, L.C., Chen, C.H., Chen, B.H., Chitrala, K.N., Chiu, Y.F., de Haan, H.G., Delgado, G.E., Demirkan, A., Duan, Q., Engmann, J., Fatumo, S.A., Gayán, J., Giulianini, F., Gong, J.H., Gustafsson, S., Hai, Y., Hartwig, F.P., He, J., Heianza, Y., Huang, T., Huerta-Chagoya, A., Hwang, M.Y., Jensen, R.A., Kawaguchi, T., Kentistou, K.A., Kim, Y.J., Kleber, M.E., Kooner, I.K., Lai, S., Lange, L.A., Langefeld, C.D., Lauzon, M., Li, M., Ligthart, S., Liu, Jun, Loh, M., Long, J., Lyssenko, V., Mangino, M., Marzi, C., Montasser, M.E., Nag, A., Nakatochi, M., Noce, D., Noordam, R., Pistis, G., Preuss, M., Raffield, L., Rasmussen-Torvik, L.J., Rich, S.S., Robertson, N.R., Rueedi, R., Ryan, K., Sanna, S., Saxena, R., Schraut, K.E., Sennblad, B., Setoh, K., Smith, A. V., Sparsø, T., Strawbridge, R.J., Takeuchi, F., Tan, J., Trompet, S., van den Akker, E., van der Most, P.J., Verweij, N., Vogel, M., Wang, H., Wang, C., Wang, N., Warren, H.R., Wen, W., Wilsgaard, T., Wong, A., Wood, A.R., Xie, T., Zafarmand, M.H., Zhao, J.H., Zhao, W., Amin, N., Arzumanyan, Z., Astrup, A., Bakker, S.J.L., Baldassarre, D., Beekman, M., Bergman, R.N., Bertoni, A., Blüher, M., Bonnycastle, L.L., Bornstein, S.R., Bowden, D.W., Cai, Q., Campbell, A., Campbell, H., Chang, Y.C., de Geus, E.J.C., Dehghan, A., Du, S., Eiriksdottir, G., Farmaki, A.E., Frånberg, M., Fuchsberger, C., Gao, Y., Gjesing, A.P., Goel, A., Han, S., Hartman, C.A., Herder, C., Hicks, A.A., Hsieh, C.H., Hsueh, W.A., Ichihara, S., Igase, M., Ikram, M.A., Johnson, W.C., Jørgensen, M.E., Joshi, P.K., Kalyani, R.R., Kandeel, F.R., Katsuya, T., Khor, C.C., Kiess, W., Kolcic, I., Kuulasmaa, T., Kuusisto, J., Läll, K., Lam, K., Lawlor, D.A., Lee, N.R., Lemaitre, R.N., Li, Honglan, Lin, S.Y., Lindström, J., Linneberg, A., Liu, Jianjun, Lorenzo, C., Matsubara, T., Matsuda, F., Mingrone, G., Mooijaart, S., Moon, S., Nabika, T., Nadkarni, G.N., Nadler, J.L., Nelis, M., Neville, M.J., Norris, J.M., Ohyagi, Y., Peters, A., Peyser, P.A., Polasek, O., Qi, Q., Raven, D., Reilly, D.F., Reiner, A., Rivideneira, F., Roll, K., Rudan, I., Sabanayagam, C., Sandow, K., Sattar, N., Schürmann, A., Shi, J., Stringham, H.M., Taylor, K.D., Teslovich, T.M., Thuesen, B., Timmers, P.R.H.J., Tremoli, E., Tsai, M.Y., Uitterlinden, A., van Dam, R.M., van Heemst, D., van Hylckama Vlieg, A., van Vliet-Ostaptchouk, J. V., Vangipurapu, J., Vestergaard, H., Wang, T., Willems van Dijk, K., Zemunik, T., Abecasis, G.R., Adair, L.S., Aguilar-Salinas, C.A., Alarcón-Riquelme, M.E., An, P., Aviles-Santa, L., Becker, D.M., Beilin, L.J., Bergmann, S., Bisgaard, H., Black, C., Boehnke, M., Boerwinkle, E., Böhm, B.O., Bønnelykke, K., Boomsma, D.I., Bottinger, E.P., Buchanan, T.A., Canouil, M., Caulfield, M.J., Chambers, J.C., Chasman, D.I., Chen, Y.D.I., Cheng, C.Y., Collins, F.S., Correa, A., Cucca, F., de Silva, H.J., Dedoussis, G., Elmståhl, S., Evans, M.K., Ferrannini, E., Ferrucci, L., Florez, J.C., Franks, P.W., Frayling, T.M., Froguel, P., Gigante, B., Goodarzi, M.O., Gordon-Larsen, P., Grallert, H., Grarup, N., Grimsgaard, S., Groop, L., Gudnason, V., Guo, X., Hamsten, A., Hansen, T., Hayward, C., Heckbert, S.R., Horta, B.L., Huang, W., Ingelsson, E., James, P.S., Jarvelin, M.R., Jonas, J.B., Jukema, J.W., Kaleebu, P., Kaplan, R., Kardia, S.L.R., Kato, N., Keinanen-Kiukaanniemi, S.M., Kim, B.J., Kivimaki, M., Koistinen, H.A., Kooner, J.S., Körner, A., Kovacs, P., Kuh, D., Kumari, M., Kutalik, Z., Laakso, M., Lakka, T.A., Launer, L.J., Leander, K., Li, Huaixing, Lin, X., Lind, L., Lindgren, C., Liu, S., Loos, R.J.F., Magnusson, P.K.E., Mahajan, A., Metspalu, A., Mook-Kanamori, D.O., Mori, T.A., Munroe, P.B., Njølstad, I., O’Connell, J.R., Oldehinkel, A.J., Ong, K.K., Padmanabhan, S., Palmer, C.N.A., Palmer, N.D., Pedersen, O., Pennell, C.E., Porteous, D.J., Pramstaller, P.P., Province, M.A., Psaty, B.M., Qi, L., Raffel, L.J., Rauramaa, R., Redline, S., Ridker, P.M., Rosendaal, F.R., Saaristo, T.E., Sandhu, M., Saramies, J., Schneiderman, N., Schwarz, P., Scott, L.J., Selvin, E., Sever, P., Shu, X. ou, Slagboom, P.E., Small, K.S., Smith, B.H., Snieder, H., Sofer, T., Sørensen, T.I.A., Spector, T.D., Stanton, A., Steves, C.J., Stumvoll, M., Sun, L., Tabara, Y., Tai, E.S., Timpson, N.J., Tönjes, A., Tuomilehto, J., Tusie, T., Uusitupa, M., van der Harst, P., van Duijn, C., Vitart, V., Vollenweider, P., Vrijkotte, T.G.M., Wagenknecht, L.E., Walker, M., Wang, Y.X., Wareham, N.J., Watanabe, R.M., Watkins, H., Wei, W.B., Wickremasinghe, A.R., Willemsen, G., Wilson, J.F., Wong, T.Y., Wu, J.Y., Xiang, A.H., Yanek, L.R., Yengo, L., Yokota, M., Zeggini, E., Zheng, W., Zonderman, A.B., Rotter, J.I., Gloyn, A.L., McCarthy, M.I., Dupuis, J., Meigs, J.B., Scott, R.A., Prokopenko, I., Leong, A., Liu, C.T., Parker, S.C.J., Mohlke, K.L., Langenberg, C., Wheeler, E., Morris, A.P., Barroso, I., de Haan, H.G., van den Akker, E., van der Most, P.J., de Geus, E.J.C., van Dam, R.M., van Heemst, D., van Hylckama Vlieg, A., van Willems van Dijk, K., de Silva, H.J., van der Harst, P., van Duijn, C., 2021. The trans-ancestral genomic architecture of glycemic traits. Nature Genetics 2021 53:6 53, 840–860.

Eastwood, S. v., Mathur, R., Atkinson, M., Brophy, S., Sudlow, C., Flaig, R., de Lusignan, S., Allen, N., Chaturvedi, N., 2016. Algorithms for the Capture and Adjudication of Prevalent and Incident Diabetes in UK Biobank. PLoS One 11, e0162388.

Elsworth, B., Lyon, M., Alexander, T., Liu, Y., Matthews, P., Hallett, J., Bates, P., Palmer, T., Haberland, V., Davey, G., 1, S., Zheng, J., Haycock, P., Gaunt, T.R., Hemani, G., 2020. The MRC IEU OpenGWAS data infrastructure. bioRxiv 2020.08.10.244293.

Gilbody, J., Borges, M.C., Smith, G.D., Sanderson, E., 2022. Multivariable MR can mitigate bias in two-sample MR using covariable-adjusted summary associations. medRxiv 2022.07.19.22277803.

Liu, M., Jiang, Y., Wedow, R., Li, Y., Brazel, D.M., Chen, F., Datta, G., Davila-Velderrain, J., McGuire, D., Tian, C., Zhan, X., Agee, M., Alipanahi, B., Auton, A., Bell, R.K., Bryc, K., Elson, S.L., Fontanillas, P., Furlotte, N.A., Hinds, D.A., Hromatka, B.S., Huber, K.E., Kleinman, A., Litterman, N.K., McIntyre, M.H., Mountain, J.L., Northover, C.A.M., Sathirapongsasuti, J.F., Sazonova, O. V., Shelton, J.F., Shringarpure, S., Tung, J.Y., Vacic, V., Wilson, C.H., Pitts, S.J., Mitchell, A., Skogholt, A.H., Winsvold, B.S., Sivertsen, B., Stordal, E., Morken, G., Kallestad, H., Heuch, I., Zwart, J.A., Fjukstad, K.K., Pedersen, L.M., Gabrielsen, M.E., Johnsen, M.B., Skrove, M., Indredavik, M.S., Drange, O.K., Bjerkeset, O., Børte, S., Stensland, S.Ø., Choquet, H., Docherty, A.R., Faul, J.D., Foerster, J.R., Fritsche, L.G., Gordon, S.D., Haessler, J., Hottenga, J.J., Huang, H., Jang, S.K., Jansen, P.R., Ling, Y., Mägi, R., Matoba, N., McMahon, G., Mulas, A., Orrù, V., Palviainen, T., Pandit, A., Reginsson, G.W., Smith, J.A., Taylor, A.E., Turman, C., Willemsen, G., Young, H., Young, K.A., Zajac, G.J.M., Zhao, W., Zhou, W., Bjornsdottir, G., Boardman, J.D., Boehnke, M., Boomsma, D.I., Chen, C., Cucca, F., Davies, G.E., Eaton, C.B., Ehringer, M.A., Esko, T., Fiorillo, E., Gillespie, N.A., Gudbjartsson, D.F., Haller, T., Harris, K.M., Heath, A.C., Hewitt, J.K., Hickie, I.B., Hokanson, J.E., Hopfer, C.J., Hunter, D.J., Iacono, W.G., Johnson, E.O., Kamatani, Y., Kardia, S.L.R., Keller, M.C., Kellis, M., Kooperberg, C., Kraft, P., Krauter, K.S., Laakso, M., Lind, P.A., Loukola, A., Lutz, S.M., Madden, P.A.F., Martin, N.G., McGue, M., McQueen, M.B., Medland, S.E., Metspalu, A., Mohlke, K.L., Nielsen, J.B., Okada, Y., Peters, U., Polderman, T.J.C., Posthuma, D., Reiner, A.P., Rice, J.P., Rimm, E., Rose, R.J., Runarsdottir, V., Stallings, M.C., Stančáková, A., Stefansson, H., Thai, K.K., Tindle, H.A., Tyrfingsson, T., Wall, T.L., Weir, D.R., Weisner, C., Whitfield, J.B., Yin, J., Zuccolo, L., Bierut, L.J., Hveem, K., Lee, J.J., Munafò, M.R., Saccone, N.L., Willer, C.J., Cornelis, M.C., David, S.P., Jorgenson, E., Kaprio, J., Stitzel, J.A., Stefansson, K., Thorgeirsson, T.E., Abecasis, G., Liu, D.J., Vrieze, S., 2019. Association studies of up to 1.2 million individuals yield new insights into the genetic etiology of tobacco and alcohol use. Nat Genet 51, 237–244.

Manichaikul, A., Mychaleckyj, J.C., Rich, S.S., Daly, K., Sale, M., Chen, W.M., 2010. Robust relationship inference in genome-wide association studies. Bioinformatics 26, 2867–2873.

Mitchell, R., Hemani, G., Dudding, T., Corbin, L., Harrison, S., Paternoster, L., 2019. UK Biobank Genetic Data: MRC-IEU Quality Control, Version 2.

Sanderson, E., Spiller, W., Bowden, J., 2021. Testing and correcting for weak and pleiotropic instruments in two-sample multivariable Mendelian randomization. Stat Med 40, 5434–5452.

Vujkovic, M., Keaton, J.M., Lynch, J.A., Miller, D.R., Zhou, J., Tcheandjieu, C., Huffman, J.E., Assimes, T.L., Lorenz, K., Zhu, X., Hilliard, A.T., Judy, R.L., Huang, J., Lee, K.M., Klarin, D., Pyarajan, S., Danesh, J., Melander, O., Rasheed, A., Mallick, N.H., Hameed, S., Qureshi, I.H., Afzal, M.N., Malik, U., Jalal, A., Abbas, S., Sheng, X., Gao, L., Kaestner, K.H., Susztak, K., Sun, Y. V., DuVall, S.L., Cho, K., Lee, J.S., Gaziano, J.M., Phillips, L.S., Meigs, J.B., Reaven, P.D., Wilson, P.W., Edwards, T.L., Rader, D.J., Damrauer, S.M., O’Donnell, C.J., Tsao, P.S., Atkinson, M.A., Powers, A.C., Naji, A., Kaestner, K.H., Abecasis, G.R., Baras, A., Cantor, M.N., Coppola, G., Economides, A.N., Lotta, L.A., Overton, J.D., Reid, J.G., Shuldiner, A.R., Beechert, C., Forsythe, C., Fuller, E.D., Gu, Z., Lattari, M., Lopez, A.E., Schleicher, T.D., Padilla, M.S., Toledo, K., Widom, L., Wolf, S.E., Pradhan, M., Manoochehri, K., Ulloa, R.H., Bai, X., Balasubramanian, S., Barnard, L., Blumenfeld, A.L., Eom, G., Habegger, L., Hawes, A., Khalid, S., Maxwell, E.K., Salerno, W.J., Staples, J.C., Yadav, A., Jones, M.B., Mitnaul, L.J., Aguayo, S.M., Ahuja, S.K., Ballas, Z.K., Bhushan, S., Boyko, E.J., Cohen, D.M., Concato, J., Constans, J.I., Dellitalia, L.J., Fayad, J.M., Fernando, R.S., Florez, H.J., Gaddy, M.A., Gappy, S.S., Gibson, G., Godschalk, M., Greco, J.A., Gupta, S., Gutierrez, S., Hammer, K.D., Hamner, M.B., Harley, J.B., Hung, A.M., Huq, M., Hurley, R.A., Iruvanti, P.R., Ivins, D.J., Jacono, F.J., Jhala, D.N., Kaminsky, L.S., Kinlay, S., Klein, J.B., Liangpunsakul, S., Lichy, J.H., Mastorides, S.M., Mathew, R.O., Mattocks, K.M., McArdle, R., Meyer, P.N., Meyer, L.J., Moorman, J.P., Morgan, T.R., Murdoch, M., Nguyen, X.M.T., Okusaga, O.O., Oursler, K.A.K., Ratcliffe, N.R., Rauchman, M.I., Robey, R.B., Ross, G.W., Servatius, R.J., Sharma, S.C., Sherman, S.E., Sonel, E., Sriram, P., Stapley, T., Striker, R.T., Tandon, N., Villareal, G., Wallbom, A.S., Wells, J.M., Whittle, J.C., Whooley, M.A., Xu, J., Yeh, S.S., Aslan, M., Brewer, J. V., Brophy, M.T., Connor, T., Argyres, D.P., Do, N. V., Hauser, E.R., Humphries, D.E., Selva, L.E., Shayan, S., Stephens, B., Whitbourne, S.B., Zhao, H., Moser, J., Beckham, J.C., Breeling, J.L., Romero, J.P.C., Huang, G.D., Ramoni, R.B., Pyarajan, S., Sun, Y. V., Cho, K., Wilson, P.W., O’Donnell, C.J., Tsao, P.S., Chang, K.M., Gaziano, J.M., Muralidhar, S., Chang, K.M., Voight, B.F., Saleheen, D., 2020. Discovery of 318 new risk loci for type 2 diabetes and related vascular outcomes among 1.4 million participants in a multi-ancestry meta-analysis. Nature Genetics 2020 52:7 52, 680–691.

Wootton, R.E., Richmond, R.C., Stuijfzand, B.G., Lawn, R.B., Sallis, H.M., Taylor, G.M.J., Hemani, G., Jones, H.J., Zammit, S., Davey Smith, G., Munafò, M.R., 2019. Evidence for causal effects of lifetime smoking on risk for depression and schizophrenia: a Mendelian randomisation study. Psychol Med 1–9.
